# Sex Differences in the Impact of Allosensitization on Waitlist Access and Post-Transplant Outcomes in Adults with Congenital Heart Disease

**DOI:** 10.64898/2026.08.31.26361832

**Authors:** Alysha Joseph, Katherine Kearney, Christine Henricks, Jamie Morgan, Weiyi Tan, Keri Shafer, Christopher Wrobel, Chantale Lacelle, Kevin Burns, Anas Jawaid, Natalie Tapaskar, Ashley Solmonson, David Nelson, Lauren K. Truby

## Abstract

**Background:** Adult congenital heart disease (ACHD) patients are prone to HLA-antibody formation from multiple surgeries, transfusions, and prosthetic surgical material. Females with ACHD may accrue additional, non-surgical alloantigen exposure. Whether sex modifies the impact of allosensitization on heart transplant (HT) access and outcomes in ACHD remains unknown.

**Methods:** We retrospectively analyzed the OPTN/UNOS registry of adults with ACHD listed for first-time HT (2018-2025). Sensitization was defined by calculated panel reactive antibodies (cPRA) at listing. We tested the sex × sensitization (highly sensitized, cPRA >50%) interaction on transplant access using Fine-Gray competing-risks regression, treating transplantation as the event of interest and death or removal from the waitlist as competing events, and on post-transplant survival using multivariable Cox proportional-hazards regression, both adjusted for age at listing, mechanical support at listing, and the number of distinct prior cardiac surgery categories.

**Results:** Among 856 candidates (38% female), females were more often highly sensitized than males (23% vs 14%; age-adjusted OR 1.81, 95% CI 1.26-2.61), even after adjusting for surgical burden. Sensitization reduced transplant access in females (84% to 71%; median wait 60 to 110 days, p < 0.001) but not males (79% vs 79%, median wait 88 vs 98 days). In adjusted Fine-Gray models, the subdistribution hazard for transplant was reduced in sensitized females (sHR 0.54, 95% CI 0.41-0.72) with no effect in males (sHR 0.96, 95% CI 0.73-1.26), and the sex × sensitization interaction was significant (interaction sHR 0.64, 95% CI 0.44-0.94, p = 0.02). Post-transplant mortality was numerically higher in sensitized than non-sensitized candidates in both sexes and the sex × sensitization interaction on 1-year mortality was not significant. The sex-asymmetric effect persisted and was more pronounced in the multiorgan candidates.

**Conclusions:** Allosensitization is not a sex-neutral barrier to transplant in HT candidates with ACHD. Females are more sensitized and have reduced transplant access without differences in 1-year mortality. The female excess in sensitization is not accounted for by surgical burden, and the exposures responsible remain to be defined. These findings warrant a sex-aware listing strategy and further studies.

## Introduction

Advances in surgical palliation and longitudinal care have resulted in a growing population of adults living with congenital heart disease (ACHD), the majority of whom now survive well into adulthood[1]. Late failure of the palliated circulation is common and often characterized by progressive ventricular dysfunction and refractory arrhythmia. Heart transplantation (HT) remains the only definitive therapy for most of these individuals with end-stage heart disease[2, 3]. As a result, adults with ACHD represent an increasing share of the transplant waitlist[2]. Unfortunately, HT candidates with ACHD continue to face disadvantages under contemporary allocation frameworks, including lower rates of HT and higher waitlist mortality[3, 4].

Allosensitization - the presence of antibodies against foreign human leukocyte antigens (HLA) - is a critical barrier to HT[5]. In the UNOS/OPTN transplant registry, the breadth of candidate sensitization is quantified as the calculated panel reactive antibody (cPRA), which estimates the proportion of donors in the general population against which a candidate carries unacceptable antigens[6]. Higher cPRA narrows the pool of immunologically compatible donors, prolongs waiting time, and is associated with worse waitlist outcomes. Sensitized candidates also face an elevated risk of antibody-mediated rejection after transplantation[7]. ACHD HT candidates are known to carry a higher burden of sensitization than other HT candidates due to multiple cardiac surgeries, blood-product transfusions, and prosthetic surgical material, particularly with cryopreserved homograft tissue used in surgical repairs of congenital lesions[8]. In ACHD HT candidates, sensitization also predicts worse post-transplant outcomes[9].

Sex differences in access to, and outcomes after HT, have been well described[10, 11], and although the 2018 allocation change improved transplantation rates for females overall, disparities have persisted under the revised system[12, 13].

Whether sex-specific sensitization contributes to these differences specifically within the ACHD population, where the baseline burden of alloantigen exposure is already high, has not been established. We therefore used the UNOS/OPTN registry to quantify and characterize sex differences in allosensitization among adults with ACHD listed for HT, and to determine whether the associations between sensitization and waitlist and post-transplant outcomes differed by sex.

## Materials and Methods

We performed a retrospective cohort analysis of the UNOS/OPTN registry to identify adults (18 years and older at listing) with ACHD listed for first-time HT from 2018-2025. Multiorgan transplant candidates were excluded from the primary analysis but analyzed as a separate cohort. Sensitization was defined by cPRA at the time of HT listing, with a cPRA of >50% being considered ‘highly sensitized’. Surgical burden was defined by the number of distinct prior cardiac surgery types (0,1, or greater than 2), derived from surgery-type coding. All patients included in the primary and secondary analysis were listed after the Heart Allocation Policy change October 18, 2018.

The primary pre-transplant outcome was access to transplant, analyzed in a competing-risks framework with time origin at the date of listing. Transplantation was the event of interest; death on the waitlist and removal from the waitlist for clinical deterioration or other reasons were treated as competing events; candidates still awaiting transplant were censored at the end of follow-up. Cumulative incidence functions for transplantation were estimated non-parametrically, and covariate effects were estimated with Fine-Gray subdistribution hazard models, implemented by constructing the finegray case-weighted risk set and fitting the corresponding weighted Cox model in the R survival package. A subdistribution hazard ratio (sHR) below 1 therefore indicates a lower rate of reaching transplant, accounting for the competing risks of death and delisting, rather than a cause-specific hazard among those remaining at risk.

We first tested the interaction between sex and high-level sensitization (cPRA >50%) on transplant access by fitting a single Fine-Gray model containing a multiplicative sex × sensitization term, and additionally fit sex-stratified models to allow the interaction to be read directly as the sensitization effect within each sex. All models were adjusted for age at listing, mechanical support at listing, and the graded number of distinct prior cardiac surgery categories. We then modeled cPRA as a continuous exposure, scaled per 10 units, and tested the sex × cPRA slope interaction. To relax the linearity assumption, cPRA was additionally modeled with a natural cubic spline (3 degrees of freedom); because the spline basis is not carried through the case-weight expansion, basis columns were precomputed and entered as ordinary covariates in both the risk-set expansion and the weighted Cox fit. Departure from linearity was assessed within each sex by a joint Wald test of the spline coefficients. Adjusted female-to-male transplant rate ratios at fixed cPRA values (0, 25, 50, 75, and 100) were obtained as linear contrasts of the interaction model coefficients, with standard errors and 95% confidence intervals derived by the delta method.

The primary post-transplant outcome was 1-year mortality, analyzed using multivariable Cox proportional hazards regression, adjusted for the same covariates, with follow-up administratively censored at 365 days. Two pre-specified sensitivity analyses were performed: restriction to blood group O candidates, and a parallel analysis of the multiorgan cohort. Descriptive comparisons used Wilcoxon rank-sum and Pearson’s chi-squared tests as appropriate, with Fisher’s exact test for sparse cells. All analyses were performed in R version 4.3.1 (R Foundation for Statistical Computing, Vienna, Austria). A two-sided P<0.05 defined statistical significance for pre-specified comparisons. The data reported here have been supplied by the Organ Procurement and Transplantation Network (OPTN). The interpretation and reporting of these data are the responsibility of the author(s) and should in no way be seen as an official policy or interpretation of the OPTN or the U.S. Government.

## Results

Among 856 ACHD HT candidates listed during the study period, 38% were female (**Table 1**). Males and females were similar in age at listing, listing era, and prior congenital surgery. MCS at listing was less common in female patients (6.5% vs 11%). cPRA data were available in 810 candidates (95%) (**Figure 1**). Females were more frequently highly sensitized than males (23% vs 14%; age-adjusted OR 1.81, 95% CI 1.26-2.61), even after adjusting for surgical burden (surgery-adjusted OR: 1.92, 95% CI: 1.33-2.78). Surgical burden was similarly distributed between sexes and did not account for the sex difference in sensitization, although sensitization increased with surgical burden in both sexes. Any detectable anti-HLA antibody (cPRA > 0) was also more common in females than males (48.4% vs 35.7%), and the sex difference was concentrated in the upper part of the cPRA distribution, as the median cPRA was 0 in both sexes, but Q3 was more than twice as high in females (46% vs 17%) (**Figure 2**).

**Figure 1.**
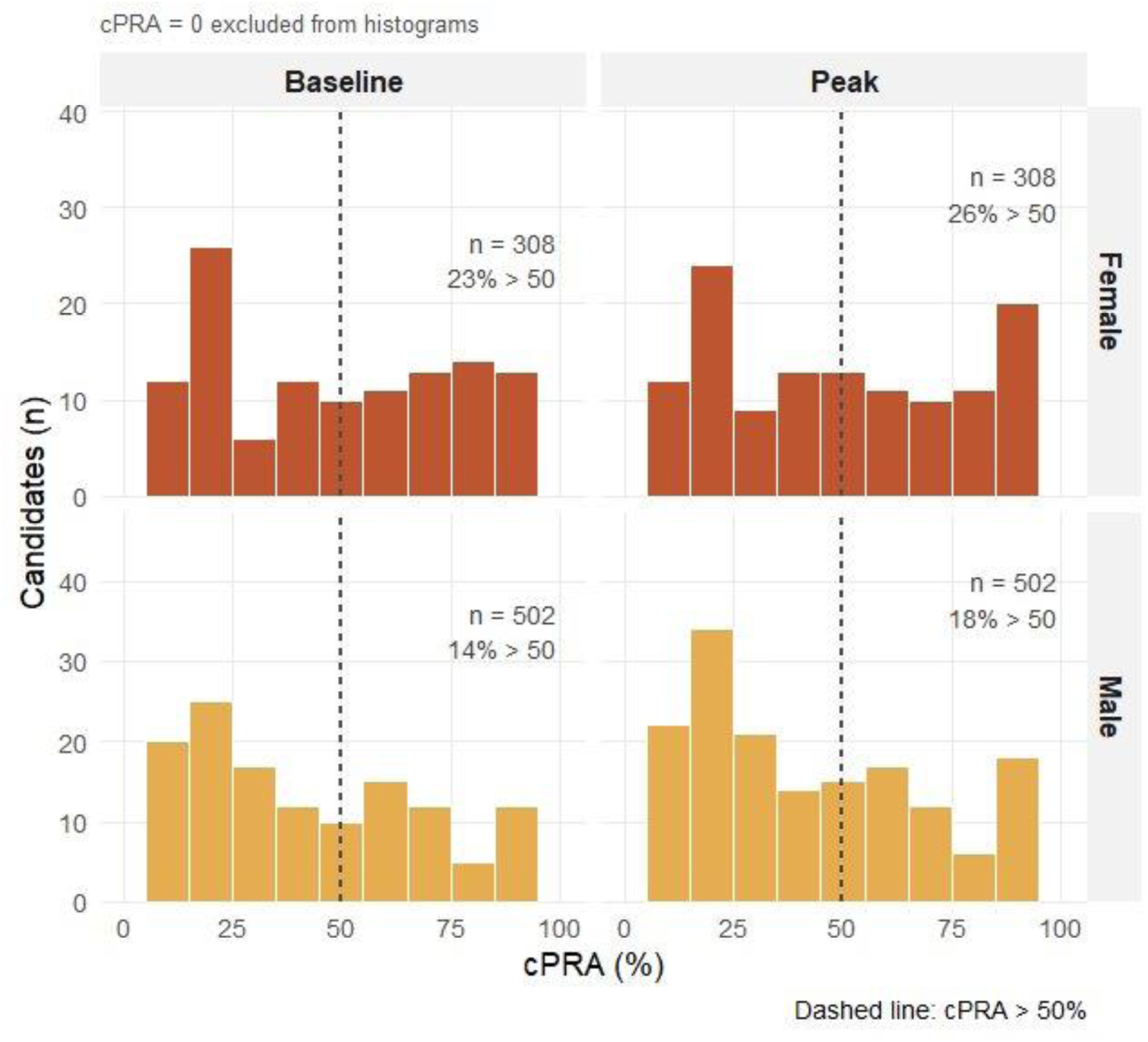
Baseline and Peak cPRA in ACHD HT Candidates by Sex.

**Figure 2.**
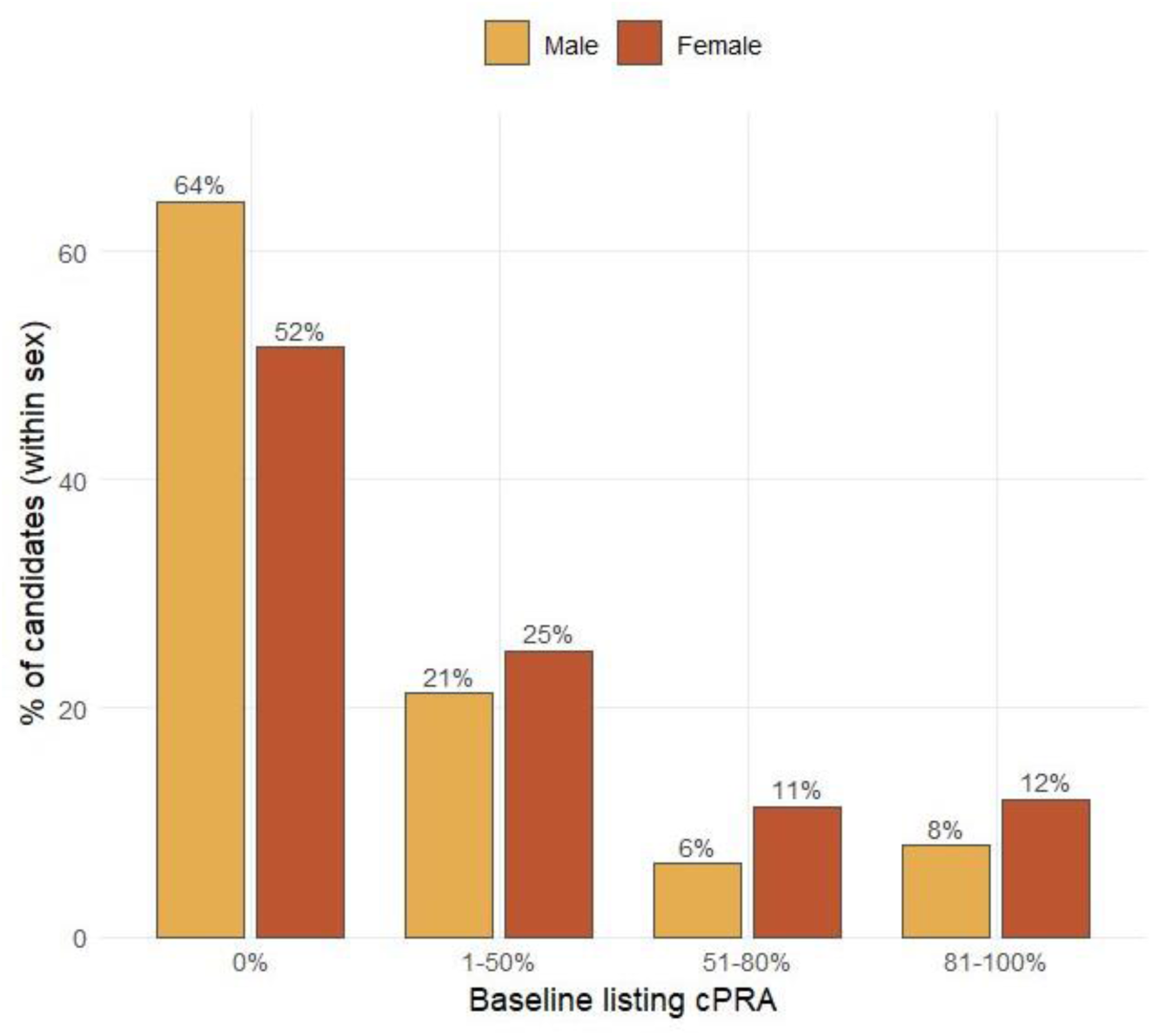
Baseline cPRA Distribution by Sex.

**Table 1.** Baseline characteristics of ACHD adults listed for first-time heart-only transplant, by sex.

| **Characteristic** | **Overall**<br>**N = 856** | **Female**<br>**N = 321** | **Male**<br>**N = 535** | **p-value** |
| --- | --- | --- | --- | --- |
| Age at listing, years | 38 (27, 48) | 36 (27, 47) | 39 (27, 49) | 0.3 |
| **Distinct prior cardiac surgery**<br>**categories** |  |  |  | **0.2** |
| 0 | 108 (13%) | 48 (15%) | 60 (11%) |  |
| 1 | 555 (66%) | 204 (65%) | 351 (67%) |  |
| >=2 | 175 (21%) | 61 (19%) | 114 (22%) |  |
| Unknown | 18 | 8 | 10 |  |
| **Prior congenital surgery** | **601 (70%)** | **217 (68%)** | **384 (72%)** | **0.2** |
| **MCS at listing** | **81 (9.5%)** | **21 (6.5%)** | **60 (11%)** | **0.024** |
| **Blood group** |  |  |  | **0.8** |
| A | 310 (36%) | 119 (37%) | 191 (36%) |  |
| AB | 26 (3.0%) | 9 (2.8%) | 17 (3.2%) |  |
| B | 121 (14%) | 41 (13%) | 80 (15%) |  |
| O | 399 (47%) | 152 (47%) | 247 (46%) |  |
| Baseline listing cPRA (%) | 0 (0, 27) | 0 (0, 46) | 0 (0, 17) | <0.001 |
| Unknown | 46 | 13 | 33 |  |
| Peak listing cPRA (%) | 0 (0, 39) | 3 (0, 52) | 0 (0, 28) | <0.001 |
| Unknown | 46 | 13 | 33 |  |
| Highly sensitized (>50%) | 144 (18%) | 72 (23%) | 72 (14%) | 0.001 |
| Unknown | 46 | 13 | 33 |  |
| Days on waitlist | 80 (21, 259) | 69 (19, 181) | 89 (22, 320) | 0.014 |
| **Waitlist outcome** |  |  |  | **0.9** |
| Died / too sick | 19 (2.2%) | 6 (1.9%) | 13 (2.4%) |  |
| Other removal | 41 (4.8%) | 14 (4.4%) | 27 (5.0%) |  |
| Still waiting | 107 (13%) | 39 (12%) | 68 (13%) |  |
| Transplanted | 689 (80%) | 262 (82%) | 427 (80%) |  |

Values are median (Q1, Q3) or n (%). p-values from Wilcoxon rank-sum test (continuous) or Pearson's chi-squared test (categorical). ACHD = adult congenital heart disease; MCS = mechanical circulatory support.

High-level sensitization was associated with substantially reduced transplant access among females but not among males with ACHD (**Figure 3**). Among heart-only-listed ACHD adults, the proportion of females transplanted fell from 84% (not highly sensitized) to 71% (highly sensitized), and median waitlist time nearly doubled (60 to 110 days, Wilcoxon p < 0.001). In males, sensitization showed no access penalty (79% vs 79%, Fisher p = 1.00; median wait 88 vs 98 days). After adjustment for age at listing, MCS at listing, and the number of distinct prior cardiac surgery categories, the Fine-Gray subdistribution hazard for transplant confirmed an estimated reduction in access among sensitized females (sHR 0.54, 95% CI 0.41–0.72, p < 0.001) with no effect in males (sHR 0.96, 95% CI 0.73–1.26, p = 0.76). The sex × sensitization interaction was statistically significant (interaction sHR 0.64, 95% CI 0.44–0.94, p = 0.02).

**Figure 3.**
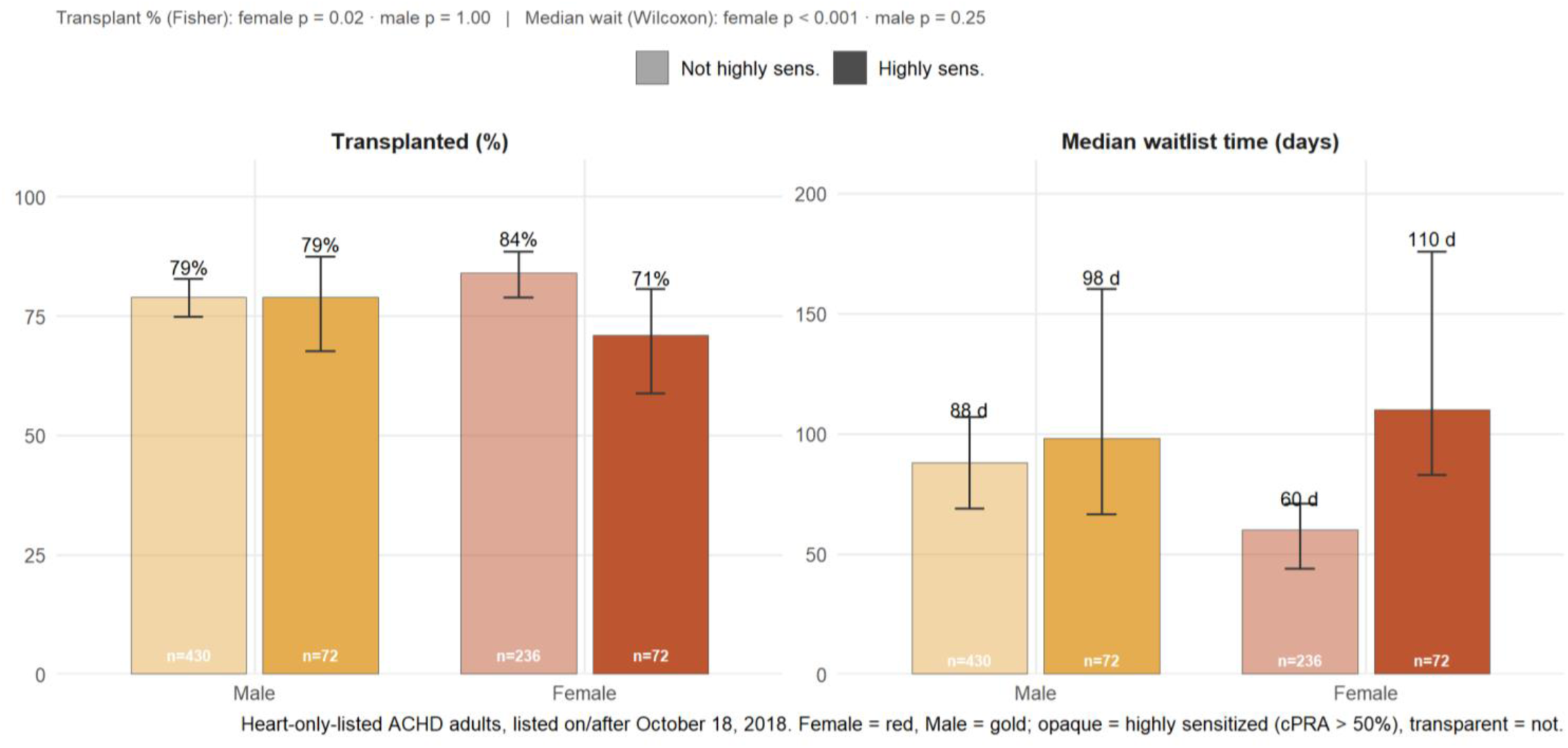
Sex-asymmetric impact of allosensitization on transplant access in ACHD.

Modeling cPRA as a continuous exposure, each 10-unit increase in baseline cPRA in females with ACHD was associated with a 7% reduction in the transplant rate (sHR 0.93 per 10 units, 95% CI 0.90–0.96, p < 0.001). The sex × cPRA slope interaction was also significant (interaction sHR 0.95 per 10 units, 95% CI 0.91–0.99, p = 0.026). The relationship between cPRA and transplant was significantly non-linear (restricted-cubic-spline joint test p < 0.001) with a monotonic decline from moderate cPRA onward, while the male curve was flat across the entire range (**Figure 4**). Between-sex comparisons at fixed cPRA values are displayed in **Table 2**. At a cPRA of 0, females reached transplant at 1.42-fold the rate of males (95% CI 1.20–1.70, p < 0.001), but this advantage narrowed to 1.10 at a cPRA of 50 (95% CI 0.91–1.34, p = 0.33) and continued to decrease to a rate ratio of 0.85 at a cPRA of 100 (95% CI 0.58–1.25, p = 0.41).

**Figure 4.**
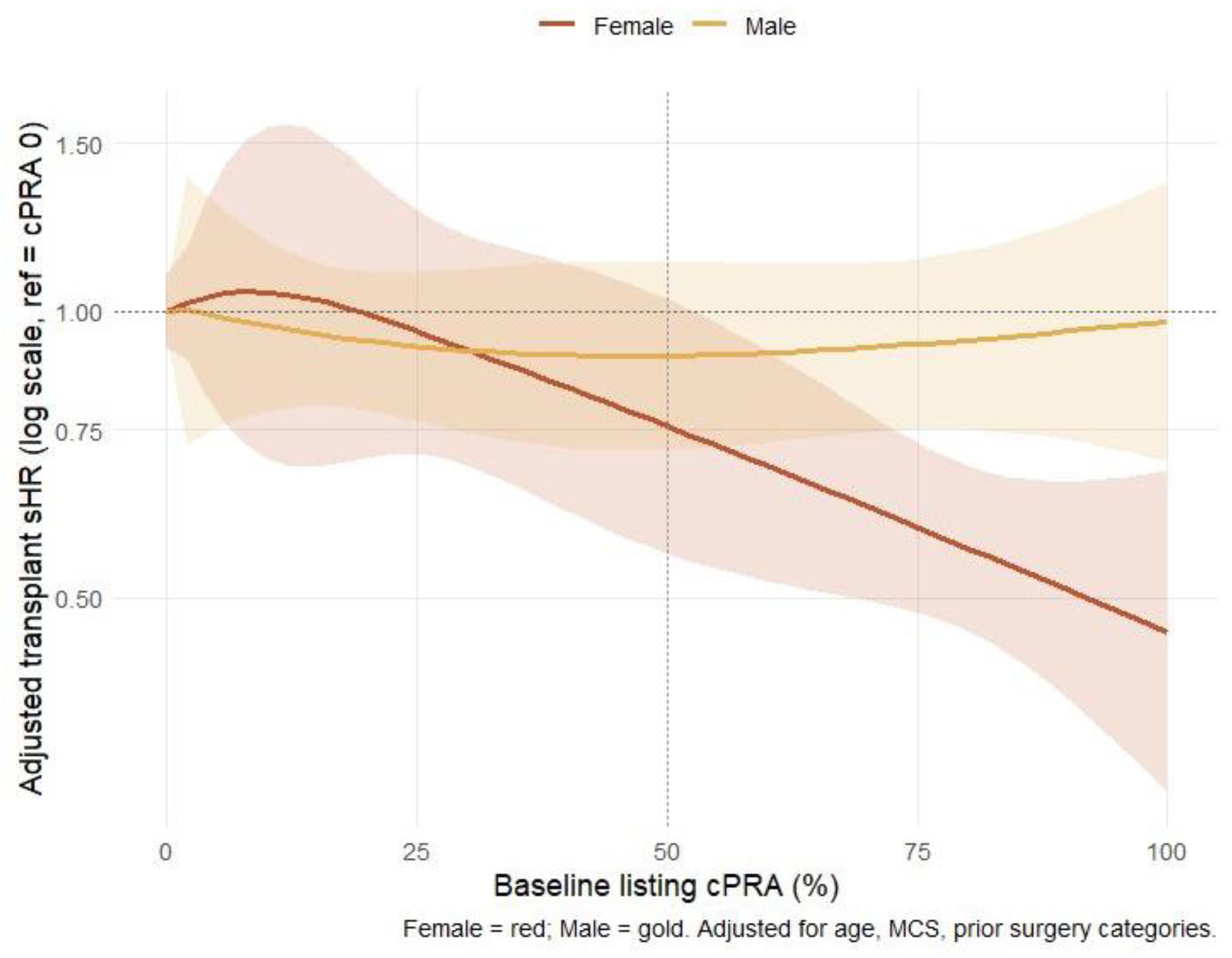
Adjusted transplant access across the cPRA range, by sex.

**Table 2:**
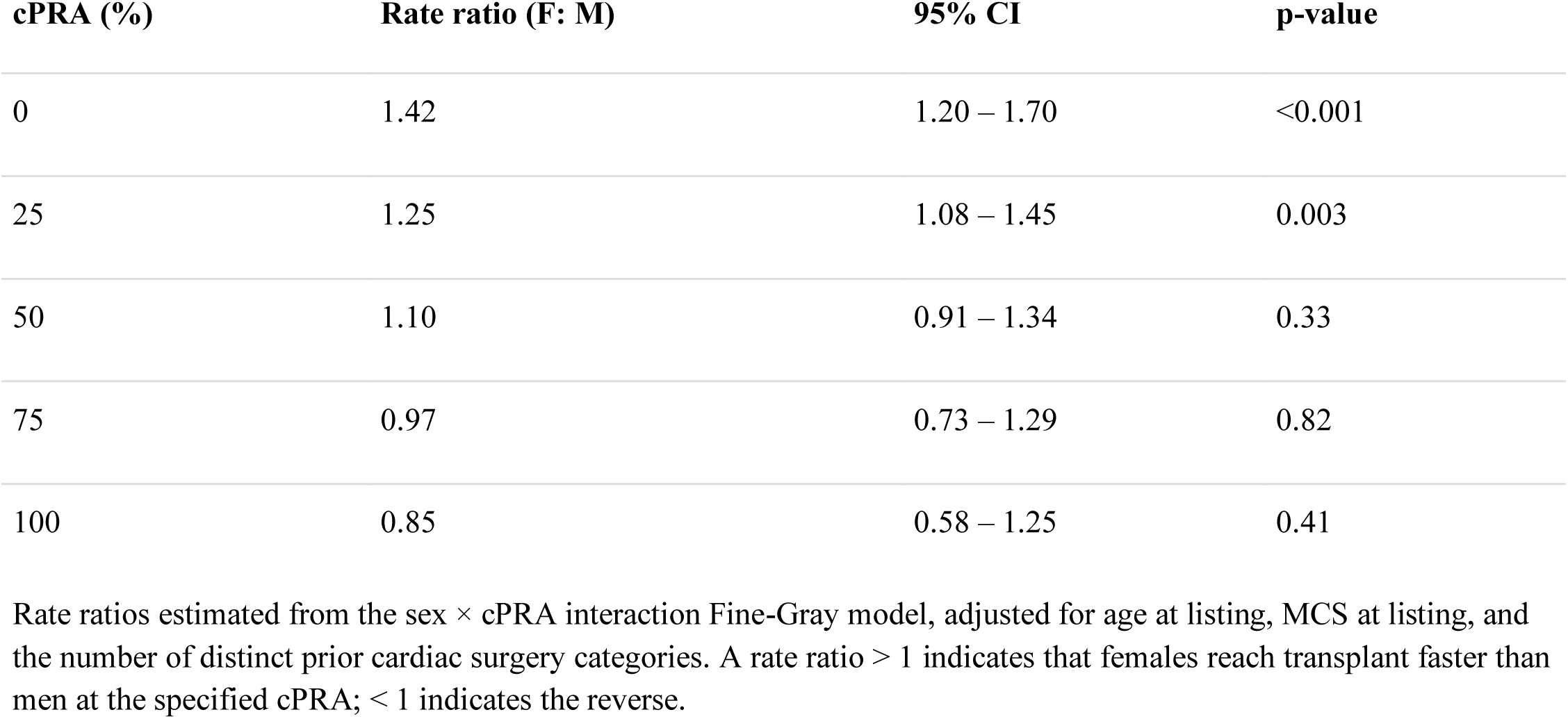
Adjusted female-to-male transplant rate ratio at fixed cPRA values.

Among the 585 transplant recipients with cPRA data (94 highly sensitized), 1-year post-transplant mortality was numerically higher in sensitized than non-sensitized candidates in both sexes: 16.7% vs 10.4% in males (9 deaths among 48 sensitized recipients; adjusted HR 1.65, 95% CI 0.75–3.67, p = 0.22) and 13.0% vs 9.8% in females (7 deaths among 46 sensitized recipients; adjusted HR 1.29, 95% CI 0.50–3.30, p = 0.59). The sex × sensitization interaction on 1-year mortality was not significant (interaction HR 0.79, 95% CI 0.23–2.70, p = 0.71) (**Figure 5**).

**Figure 5.**
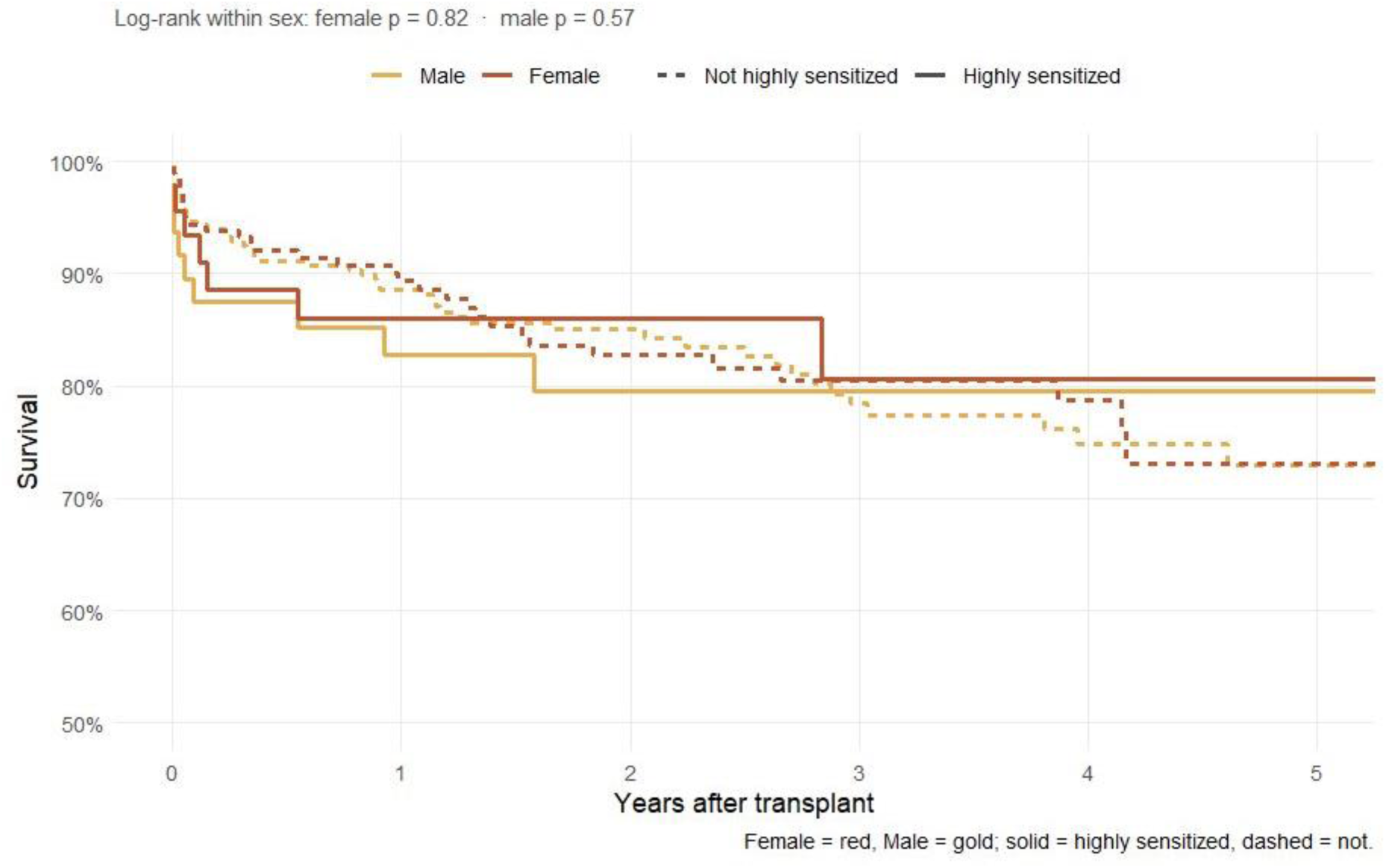
Post-transplant survival in ACHD adults, by sex and sensitization.

We performed two sensitivity analyses. The first was to understand whether blood type, specifically blood type O, impacted access across sexes. Sensitized females with blood type O incurred similar rates of transplantation as sensitized females overall (adjusted sHR 0.54, 95% CI 0.41–0.71, p < 0.001), suggesting that the female sensitization penalty is not explained by the compounded ABO-plus-HLA donor-pool restriction that group O sensitized candidates might be expected to face. The second was to understand the role of sex-specific sensitization in multiorgan candidates. Among the 308 ACHD adults listed as multi-organ candidates during the study period (130 females, 178 males; 292 with baseline cPRA), the sex-asymmetric impact of sensitization on transplant access was preserved. Sensitized multi-organ ACHD females were transplanted at only 58% (n = 31; median wait 264 days) compared with 79% of non-sensitized multi-organ ACHD females (n = 92; median wait 116 days). Sensitized multi-organ ACHD males, in contrast, showed no access penalty (84% transplanted, median wait 133 days vs 76%, 149 days in non-sensitized males). Post-transplant 1-year mortality in multi-organ recipients was substantially higher than in heart-only recipients across all sex × sensitization cells (25–46% at 1 year vs 10–17% in heart-only), reflecting the greater peri-operative and immunologic complexity of multi-organ transplantation in this population.

## Discussion

This is the first study to examine sex differences in allosensitization on waitlist and post-transplant outcomes in ACHD patients. We found that female ACHD patients were more sensitized than male patients (23% vs 14%), even after adjusting for surgical burden. High-level sensitization reduced transplant access in female but not male patients (adjusted sHR 0.54 vs 0.96, interaction p = 0.02). One-year post-transplant mortality trended higher in sensitized male patients but did not reach significance in either sex. Neither blood type (ABO O) nor multiorgan transplantation appeared to attenuate this association.

At equivalent listing cPRA, female and male ACHD patients had differential waitlist times, even after adjusting for other factors – age, transplant era, MCS, and surgical burden. In the general heart transplant population, females had equal or better access under the 2018 allocation system, transplanted at higher rates yet with modestly worse adjusted post-transplant survival[14]. In the ACHD population, with sensitization considered, the opposite relationship was observed, with lower transplant rates for sensitized females and no trend towards increased post-transplant mortality. We propose two broad categories of explanation: biological drivers rooted in the sensitizing exposure itself, and systemic decision-level drivers at the candidate-donor interface.

Pregnancy is one plausible, but untested, driver of the sex-specific access gap seen in this study. In a US multisite surveillance study of 26,655 women with congenital heart defects aged 11 to 50 years, 21.3% had diagnostic codes indicating a pregnancy over a three-year window, with age-adjusted three-year pregnancy proportions of 10.0% to 24.6% among those with severe defects and 14.2% to 21.7% among those with non-severe defects[15]. In a Norwegian nationwide cohort of women of reproductive age, 77.1% of those with mild CHD and 63.6% of those with moderate or severe CHD had become mothers by age 40, with a mean of 1.81 and 1.42 childbirths per woman, respectively, compared with 76.8% and 1.80 among women without heart disease[16]. Pregnancy exposes the maternal immune system to paternally inherited fetal antigens, leading to the development of durable anti-HLA antibodies that increase with parity [17]. Because placental trophoblast lacks classical HLA class I and II, sensitization occurs indirectly. Fetal antigens enter the maternal circulation as fetal cells, cell-free DNA, and microvesicles are presented by maternal antigen-presenting cells on MHC class II, and prime a CD4+ T cell–dependent response producing increasing anti-HLA class II antibody through the third trimester [18, 19]. The resulting antibody profile varies with obstetric history: primiparous women predominantly carry class II antibodies, while multiparous women carry both class I and II antibodies [20]. The degree of maternal-fetal HLA epitope mismatch predicts the development of child-specific antibodies, an established risk factor for donor HLA-specific antibodies after organ transplantation. [21, 22].

Pregnancy-induced anti-HLA sensitization may be distinct from surgery or transfusion-induced sensitization and carry its own antibody characteristics and rejection risk profile[23]. This matters for transplant access because class II antibodies may exert a greater impact on cPRA, due to higher antigenic diversity of class II as opposed to class I antibodies, which may translate into disproportionate cPRA points and more restricted effective donor pool for patients with pregnancy-derived antibodies as opposed to surgical or transfusion exposure [24]. Thus, pregnancy-derived sensitization may impose a steeper access penalty per unit of cPRA, offering a potential explanation for the disparity observed in highly sensitized females in this study. Partner HLA typing could improve pre-transplant risk assessment, and murine data showing distinct plasticity in pregnancy-induced memory B cells suggest possible targets for improving transplant access in this population [25, 26].

An alternative explanation for the access penalty lies in donor offer acceptance patterns. Sex asymmetry in acceptance between a sensitized female candidate with a narrow acceptable-antigen profile and an equivalent male candidate could arise from perceived crossmatch difficulty, size-matching constraints, or a lower threshold for continuing to wait [27]. A related mechanism operates upstream of listing: if providers are less likely to refer or list women they expect to be difficult to match, our estimate understates the true penalty, because a waitlist-based cohort cannot capture candidates who were never listed. This referral bias could also explain the lower proportion of female ACHD patients listed for first-time heart-only transplant in our cohort—and it means our findings, if anything, are conservative.

These drivers may coexist with a broader access–survival tradeoff at the candidate–donor interface: sensitized females may wait longer for better-matched donors, improving post-transplant survival at the cost of access, while sensitized males may be accepted with less favorable matches, trading access for survival risk [4]. Alternatively, high sensitization in males may mark a sicker population with greater exposure to sensitizing events (transfusions, homograft material), consistent with their trend toward increased mortality.

For sensitized female ACHD patients, the dominant penalty is access to transplant, without a significant impact on mortality. Several interventions could narrow this gap. Desensitization protocols reduce antibody burden and broaden the acceptable donor pool, though their durability and effect on post-transplant outcomes in ACHD remain unestablished[28]. More granular acceptable-antigen listing, rather than reliance on a single cPRA threshold, would also allow for an expanded donor pool. Allocation-based priority adjustments may help to narrow the gap in sex-differences in this population[29].

There are several limitations to this study. ACHD lesion complexity may modify sensitization and outcomes, but it is not captured in this registry. Furthermore, this registry data does not capture obstetric history; therefore, we cannot directly test whether pregnancy-related sensitization specifically drives the female access penalty. The pregnancy mechanism discussed above should be regarded as a hypothesis for future study rather than a finding of this analysis. We also lack antibody profile detail (class I vs II, antibody strength, specificity), the exact unacceptable antigens listed, and offer-level acceptance data, which are outside the scope of the study. The Fine-Gray approach models the subdistribution hazard and therefore estimates effects on the cumulative incidence of transplantation rather than on the instantaneous rate among those still at risk; cause-specific and subdistribution hazards answer different questions and should not be interpreted interchangeably. The retrospective, observational design of the study limits causal inference. These findings are hypothesis-generating.

In conclusion, ACHD female HT candidates are more sensitized and have reduced transplant access without a difference in 1-year mortality. Pregnancy may play a role in this disparity, as sensitization exceeds what surgical burden explains. These findings warrant a sex-aware listing strategy, with proactive ways to mitigate sensitization in high-risk candidates (both males and females). Pregnancy may be an opportunity to mitigate risk in females. Multidisciplinary collaboration among cardiology, maternal-fetal medicine, and immunology will be essential for future studies.

## Data Availability

All data utilized in the present study are available by request from OPTN/UNOS after execution of appropriate data use agreement.

## Non-standard abbreviations

ACHD: (Adult Congenital Heart Disease)
CPRA: (Calculated Panel-reactive Antibody)
MCS: (Mechanical Circulatory Support)
HT: (Heart Transplant)
OPTN: (Organ Procurement And Transplantation Network)
UNOS: (United Network For Organ Sharing)
HLA: (Human Leukocyte Antigen)
CHD: (Congenital Heart Disease)

